# Impact of COVID-19 on Hepatitis B Screening in Sierra Leone: Insights from a Community Pharmacy Model of Care

**DOI:** 10.1101/2023.05.24.23290418

**Authors:** Manal Ghazzawi, Lawrence S. Babawo, Amir M. Mohareb, Peter B. James, Sahr A. Yendewa, Samuel P.E. Massaquoi, Peterlyn E. Cummings, Sulaiman Lakoh, Robert A. Salata, George A. Yendewa

## Abstract

**Background:** There are limited studies evaluating the impact of COVID-19-related interruptions on hepatitis B virus (HBV) screening in endemic countries in Sub-Saharan Africa.

**Methods:** We conducted a retrospective study of HBV testing in a community pharmacy in Freetown, Sierra Leone, from October 1, 2019, through September 30, 2022. We compared participant characteristics using Pearson’s chi-square test. We evaluated trends in HBV screening and diagnosis using one-way ANOVA with Tukey’s or Dunnett’s post-test.

**Findings:** Of 920 individuals screened, 161 had detectable HBsAg (seroprevalence 17.5% [95% CI 14.9-20.4]). There was a 100% decrease in HBV screening during January-June of 2020; however, screening increased by 27% and 23% in the first and second year after COVID-19, respectively. Mean quarterly tests showed a significant upward trend: 55 ± 6 tests during January-March (baseline), 74 ± 16 tests during April-June, 101 ± 3 tests during July-September, and 107 ± 17 tests during October-December (one-way ANOVA test for trend, F=7.7, p = 0.0254) but not the mean quarterly number of people diagnosed with HBV (F = 0.34, p = 0.7992).

**Interpretation:** Community-based HBV screening dramatically improved following temporary disruptions related to COVID-19. Seasonal variation in HBV screening, but not HBV diagnosis, may have implications for HBV elimination efforts in Sierra Leone and other West African countries.

## INTRODUCTION

Hepatitis B infection (HBV) affects 296 million people globally, with Sub-Saharan Africa (SSA) accounting for 25% of chronic cases, i.e., 80 million people [1]. Without timely diagnosis and treatment, there is an increased risk of developing cirrhosis, hepatocellular carcinoma, and end-stage liver disease, which collectively caused 820,000 deaths in 2019 [1]. The World Health Organization (WHO) has set a goal of eliminating HBV globally by 2030 through a 90% reduction in incidence and a 65% decrease in mortality [2].

Despite the high global burden of HBV, progress towards the elimination goals is threatened by limited access to HBV screening [2, 3]. Awareness of HBV status is the first critical step in navigating the HBV cascade of care and facilitates linkage to essential healthcare services. However, in 2021, only 10% of people with chronic HBV (PWHB) globally were aware of their status, with an even lower proportion of just 2% in the WHO African region [4]. Furthermore, many SSA countries lack adequate resources and infrastructure for delivering affordable HBV services [4, 5], and existing pathways for accessing these services are largely reliant on hospital-or clinic-based HIV facilities [5-7]. Additionally, the coronavirus-19 (COVID-19) pandemic has had a notable impact on healthcare delivery globally and further complicated efforts to eliminate HBV, due to prioritization of the COVID-19 pandemic response [8-11].

To address the gap in affordable and accessible HBV screening, alternative models of care delivery such as community pharmacies have emerged in both high- and low-income countries [12-14]. This has been facilitated by the widespread availability of point-of-care (POC) tests. Commercially available POC tests for HBV have excellent diagnostic performance and offer an alternative to laboratory-based testing [15, 16]. The advantages of POC tests include low cost, ease of use requiring minimal training, quick results turnaround, and scalability, which increases their accessibility to hard-to-reach populations [15]. Moreover, there is evidence that nontraditional settings such as pharmacies, drug and harm reduction services, mobile health units, and community-based organizations are preferred for HIV, viral hepatitis, and sexually transmitted infections services by marginalized or stigmatized groups [17, 18], presenting an opportunity to target populations most in need of care and accelerate HBV elimination efforts.

Sierra Leone is a West African country with an estimated national prevalence of HBV surface antigen (HBsAg) of 13% [19]. While there have been notable strides to implement a national policy for combating HBV, the delivery of HBV care remains fragmented. There are few viral hepatitis specialty clinics in Sierra Leone, and HBV care delivery is primarily through hospital-based HIV services [7]. Like elsewhere, the COVID-19 pandemic has hindered healthcare delivery in the country [20-22], however, its impact on HBV care has not been evaluated.

The objectives of our study were to: (1) evaluate the impact of COVID-19 on the utilization of HBV screening services in Sierra Leone and (2) assess trends in HBV screening and diagnosis using a community-based pharmacy model of care.

## METHODS

### Study setting and context

KnowHep Foundation Sierra Leone is a local non-governmental organization committed to raising public awareness about viral hepatitis through mass education and advocacy within communities across Sierra Leone. Established in 2019, the main objective of the foundation is to improve viral hepatitis care delivery by reaching a minimum of 80% of the population of Sierra Leone by 2030, through community outreach initiatives that support the global viral hepatitis elimination agenda. KnowHep Foundation is affiliated with CitiGlobe Pharmacies Ltd, a network of community-based pharmacies that provide free or low-cost HBV screening, linkage to care for individuals who test positive for HBV, and HBV vaccination services.

### Study design and key time points

We conducted a retrospective review of pharmacy HBV testing records at CitiGlobe Pharmacy Ltd in Freetown, Sierra Leone, from October 1, 2019, when meticulous documenting of all HBV tests performed was commenced, through September 30, 2022. The first confirmed cases of COVID-19 in Sierra Leone were reported in late March 2020, however, the COVID-19 pandemic had already spread in China, Europe, and elsewhere since December 2019. This resulted in major disruptions in global supply chains for medications, testing kits, and other healthcare essentials. Additionally, there were growing concerns about COVID-19 locally in Sierra Leone, which led to fewer pharmacy visits by the public. As a result of this, pharmacy services were suspended in the first quarter of 2020. With the identification of the first COVID-19 cases in Sierra Leone in March 2020, the Government of Sierra Leone declared a state of emergency, and a series of population-wide shelter-in-place orders were imposed in the country from April through June 2020. The suspension of pharmacy services continued throughout this period. The shelter-in-place orders were lifted at the end of June 2020, which allowed the resumption of pharmacy services in July 2020.

We divided the study period into the following testing periods: (1) Year 1: October 2019-September 2020; (2) Year 2: October 2020-September 2021; and (3) Year 3: October 2021-September 2022. We designated Year 1 as the baseline year for HBV testing and compared it with Years 2 and 3. To assess seasonal trends in HBV screening, we further divided each year into quarters (i.e., three-month periods).

### Data collection and definitions

We used a structured data collection instrument to extract the following data from screening records: evidence of chronic HBV infection, defined as HBsAg detection, date of screening, sex, age, highest education attained, and testing episode, i.e., first-time screening or repeat testing for clinical monitoring of previously diagnosed infection. All personal identifiers (i.e., name, date of birth, and address) were removed before entering the data into a password-protected spreadsheet for analysis.

### Statistical analysis

We report descriptive statistics as frequencies with percentages, means with standard deviation, or medians with range. We calculated associations between variables using Pearson’s chi-square test by SPSS Version 29.0 (Armonk, NY; IBM Corp). We estimated the mean quarterly number of HBV tests as the average of the tests performed during corresponding quarters in each of Years 1, 2, and 3, respectively. We assessed seasonal trends in quarterly HBV screening and diagnosis using one-way ANOVA with Tukey’s or Dunnett’s post-test by GraphPad Prism Version 9.5.1 (San Diego, California, USA). In all analyses, results were considered statistically significant when p < 0.05.

### Ethical consideration

We obtained ethical approval to perform the study from Njala University, Bo Campus, Sierra Leone (approval date 23 January 2023). Informed consent was not required as this study involved a retrospective analysis of secondary data.

## RESULTS

### Characteristics of the study participants

A total of 1145 unique individuals underwent HBV screening at the pharmacy from October 2019 through September 2022. Of these, 920 individuals underwent HBV screening for the first time, while 225 individuals had repeat HBV testing for clinical monitoring of previously diagnosed HBV infection. Table 1 presents baseline sociodemographic characteristics of the population of interest, i.e., first-time testers only. This consisted of roughly equal proportions of males (51.7%, 476/920) and females (48.3%, 444/920). The median age was 32 (IQR 27-38) years and the majority were aged 30-39 years. We found that 52.4% (482 of 920) had attained the tertiary level of education.

**Table 1.**
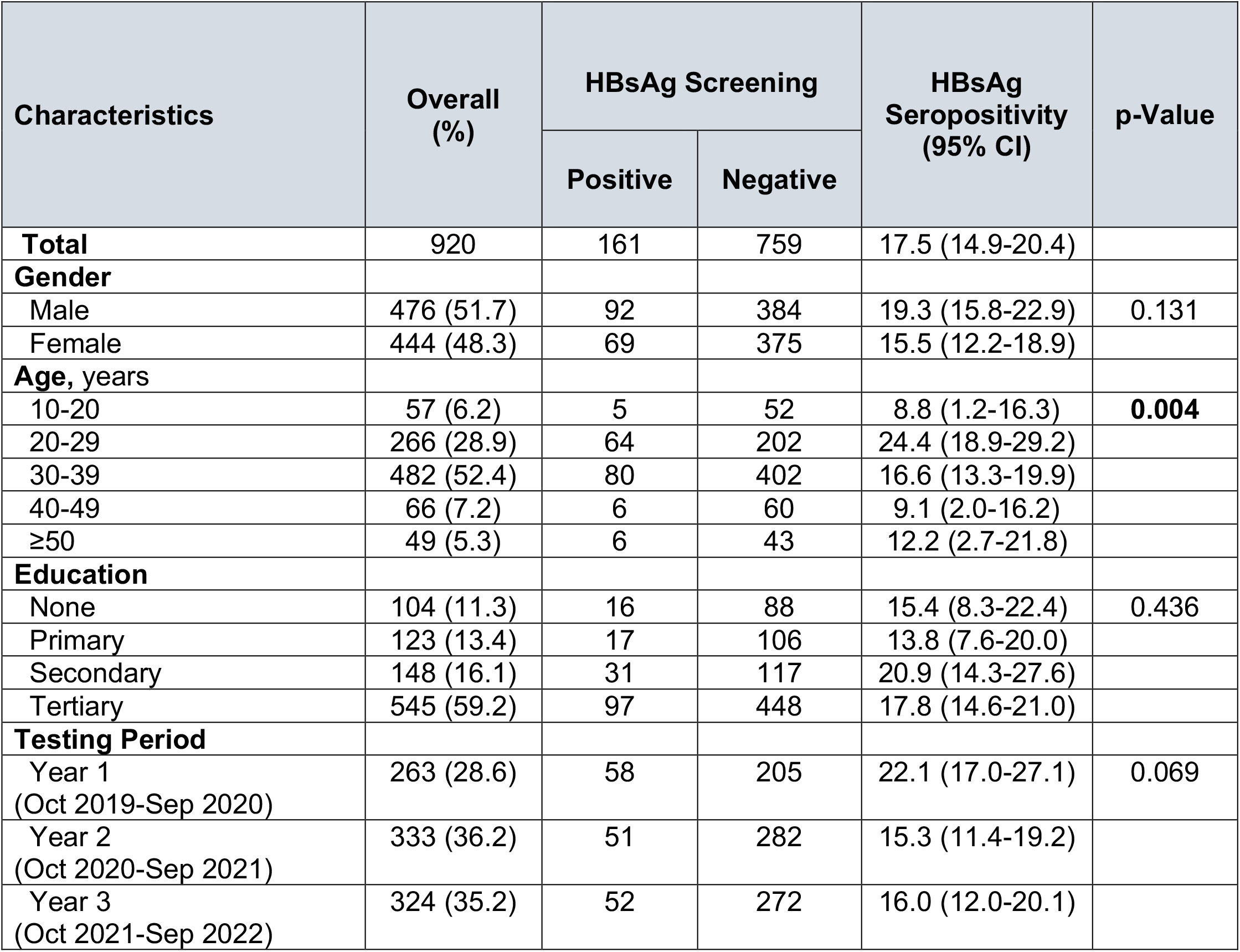
Seroprevalence rates of HBsAg by baseline characteristics and testing periods

### Seroprevalence of HBsAg

As shown in Table 1, the overall seroprevalence of HBsAg was 17.5% (95% CI 14.9-20.4). When disaggregated by sociodemographic categories, individuals aged 30-39 years had significantly higher HBsAg seropositivity compared with individuals of other age groups, i.e., 24.4% (95% 18.9-29.2); p = 0.004. The HBsAg seroprevalence did not vary based on sex, educational attainment, or year of testing.

### Impact of COVID-19 on HBV Screening

Overall, 263 individuals were screened for HBV during Year 1, compared with 333 individuals during Year 2 (i.e., a 27% increase from Year 1) and 324 individuals during Year 3 (i.e., a 23% increase from Year 1). Figure 1 presents the quarterly number of HBV tests performed, while Figure 2 presents the percentage change in quarterly screenings compared with the pre-COVID-19 pandemic baseline, i.e., October-December 2019. A total of 122 HBV tests were performed during the baseline period. Due to the COVID-19-related suspension of pharmacy services, no HBV screening was offered during January-June 2020, representing a 100% decrease from the pre-COVID-19 baseline (Figure 2). Following the lifting of the restrictions, 141 HBV tests were performed during the third quarter of 2020, representing a 16% increase compared to the pre-COVID-19 baseline. However, this increase in HBV testing was not sustained over subsequent quarters in Years 1, 2, or 3 (Figure 2).

**Figure 1.**
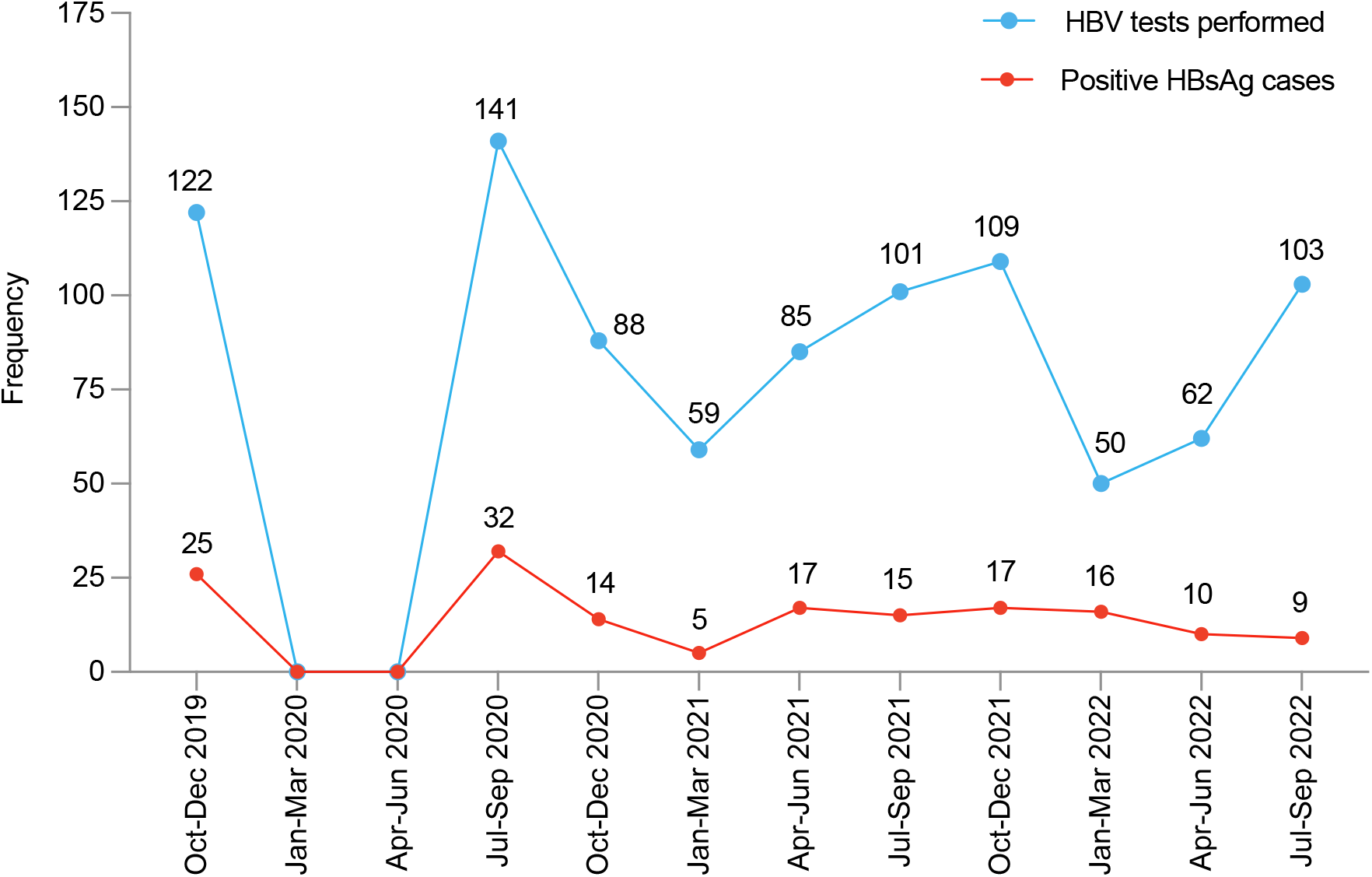
Temporal trends in HBV testing and HBsAg seroprevalence from October 2019 to September 2022 Abbreviations: HBV, hepatitis B virus; HBsAg, hepatitis B surface antigen

**Figure 2.**
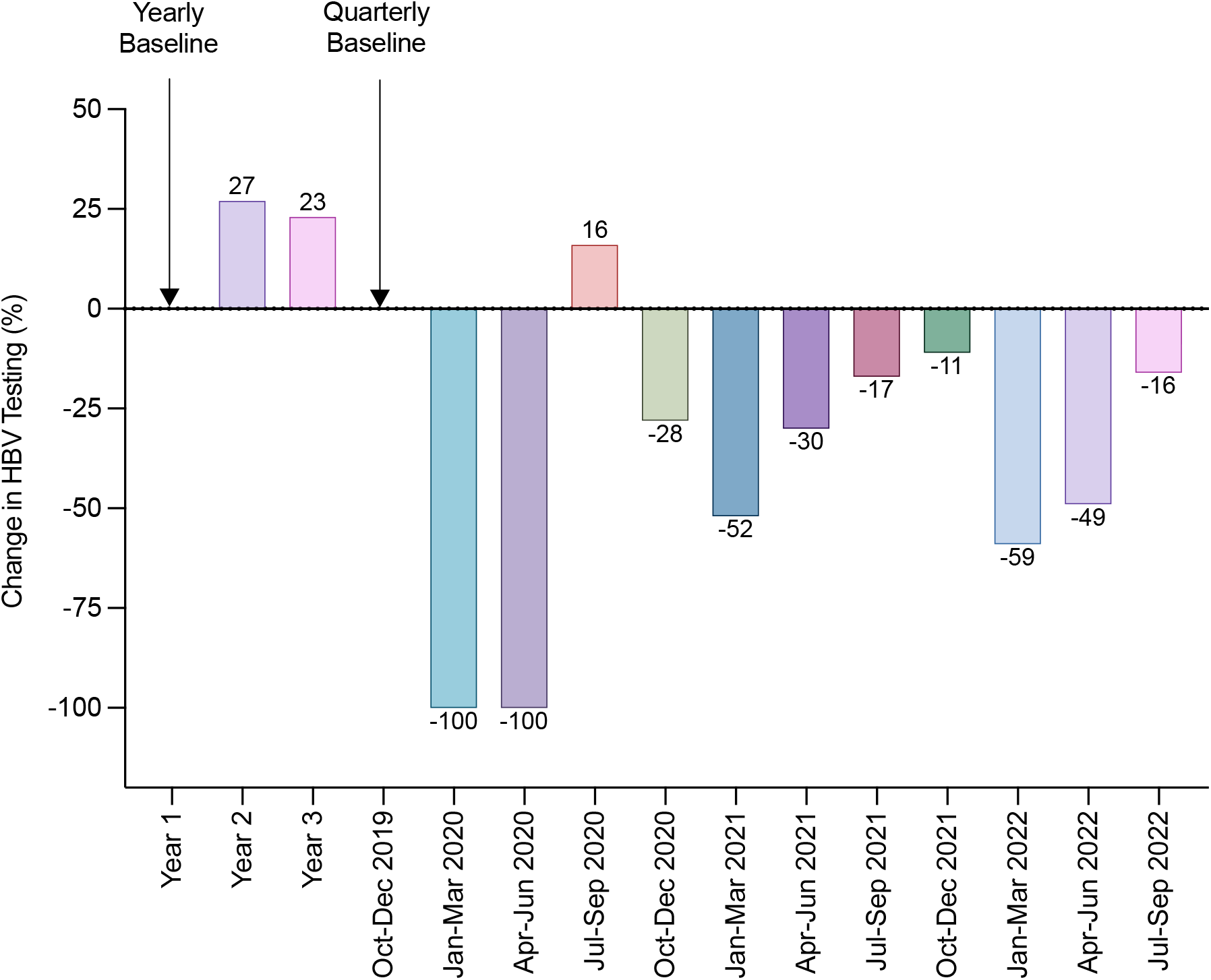
Changes in HBV screening from October 2019 to September 2022 Abbreviations: HBV, hepatitis B virus

### Seasonal Trends in HBV Screening and Diagnosis

Figure 3 presents the mean number of HBV tests performed per quarter over Years 1, 2, and 3, after excluding the first, second, and third quarters of 2020 from the analysis to account for the disruptions in testing patterns due to the COVID-19 pandemic. On average, 55 ± 6 HBV tests were performed during January-March (baseline), 74 ± 16 tests during April-June, 101 ± 3 tests during July-September, and 107 ± 17 tests during October-December. One-way ANOVA showed that there was no statistically significant difference in the number of HBV tests performed between January-March and April-June (p = 0.4237); however, there were significantly more tests performed during July-September (p = 0.0380) and October-December (p = 0.0192), respectively, compared with the January-March baseline. Test of trends between quarterly means by one-way ANOVA confirmed that HBV testing exhibited a linear seasonal trend, with the lowest number of tests performed in the first half of the year, which increased significantly throughout the remainder of the year (F = 7.7, p = 0.0254).

**Figure 3.**
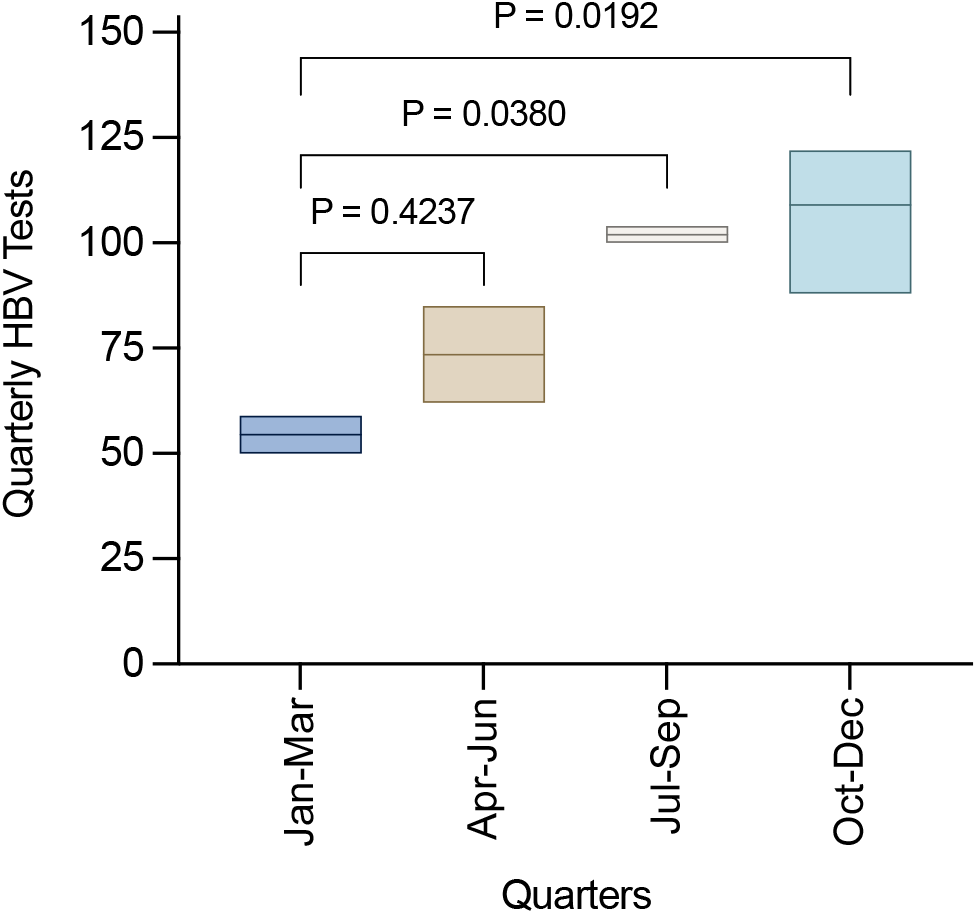
Quarterly trends in HBV testing Abbreviations: HBV, hepatitis B virus

As shown in Figure 4, the mean number of diagnosed cases, i.e., HBsAg positive tests per quarter, were 11 ± 2 during January-March, 14 ± 5 during April-June, 12 ± 4 during July-September, and 16 ± 2 during October-December. There was no significant difference in the mean quarterly number of people diagnosed with HBV, i.e., HbsAg seropositive tests (p > 0.05 for all quarters). Test for trend by one-way ANOVA did not show a trend in the mean quarterly number of diagnosed cases (F = 0.34, p = 0.7992).

**Figure 4.**
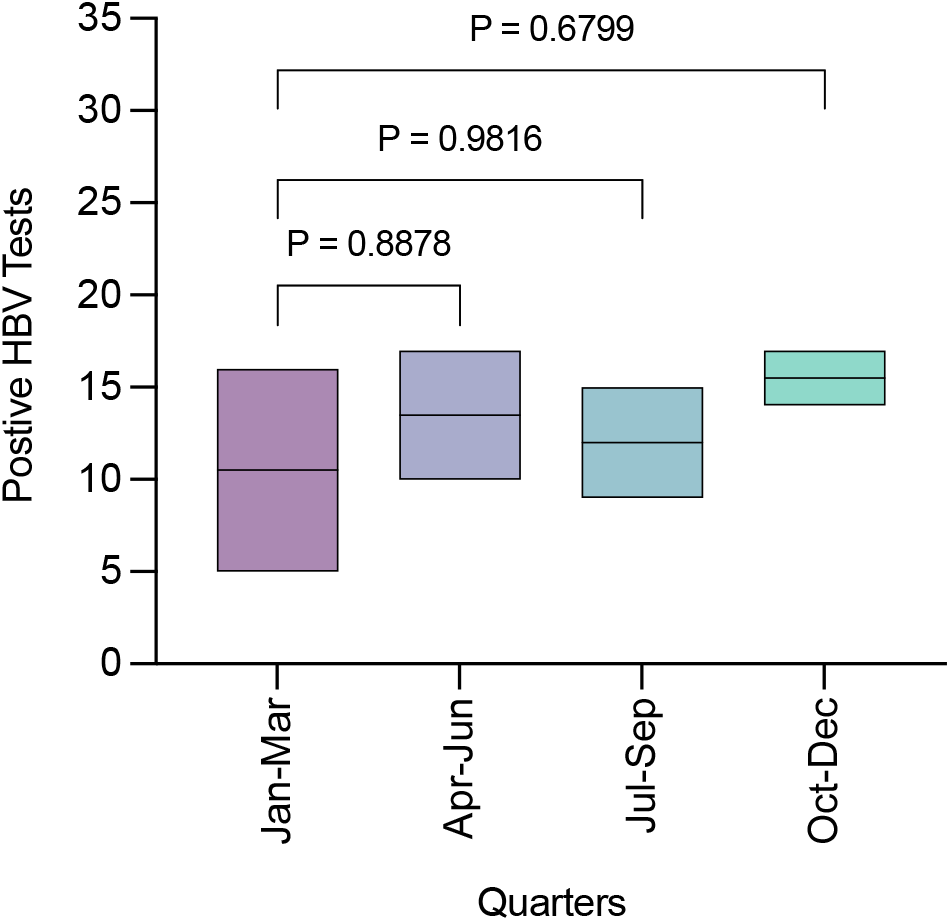
Quarterly trends in HBV diagnosis Abbreviations: HBV, hepatitis B virus

## DISCUSSION

In this study of community-based HBV screening in a pharmacy in Freetown, Sierra Leone, we demonstrate trends in HBV screening and diagnosis. A total of 920 individuals underwent first-time screening for HBV at our pharmacy over the three-year study period, of whom 17.5% tested positive for HBsAg. The first year of testing coincided with the onset of the COVID-19 pandemic, which saw major disruptions in service delivery. As a result, there was a 100% decrease in HBV testing in the first and second quarters of 2020 compared with the pre-COVID-19 baseline period. This was due to the fact that in the first quarter of 2020, pharmacy visits decreased considerably partly due to well-known seasonal variations associated with the Christmas and New Year festivities in this setting, as well as growing public concerns about the COVID-19 pandemic which had already spread in China, Europe and elsewhere since December 2019. Due to these challenges, pharmacy services were suspended in the first quarter of 2020 and remained so during the population-wide lockdowns implemented in Sierra Leone in the second quarter of 2020.

Our observations are consistent with research from both high- and low-income settings, which have reported significant disruptions in HBV service delivery resulting from the COVID-19 pandemic measures. In a major study sponsored by the European Association for the Study of the Liver, Kondili et al [10] reported a 35% reduction in HBV testing between January 2019 and December 2020 in 32 European and 12 non-European clinical centers. The impact of the COVID-19 pandemic was also observed across multiple levels of the HBV cascade of care, with a 30% reduction in consultations, a 39% decrease in referrals, a 29% decline in HBV viral load testing, and a 35% drop in treatment initiation [10]. Countries in sub-Saharan Africa have faced similar challenges in HBV care delivery. Lemoine et al [11] reported that between January and April 2020, the monthly number of visits at hepatitis clinics for new cases declined significantly by 71% in Burkina Faso, 95% in Tanzania, and 83% in The Gambia, and for patients in follow-up, the numbers fell by 73%, 77%, and 89% in Burkina Faso, Tanzania, and The Gambia, respectively. In comparison, the utilization of outpatient services declined by 18% during the West African Ebola epidemic of 2014 [23], indicating the severity of the impact of infectious disease outbreaks on routine healthcare delivery.

Despite the pandemic-related disruptions in 2020, we observed an overall 27% and 23% increase in HBV screening in our pharmacy in the second and third testing years, respectively, reflecting the healthcare system’s ability to address short-term increases in demand for services. However, it is uncertain whether this trend will continue in the future. Achieving HBV elimination targets requires a sustained increase in HBV screening, treatment and prevention, and the healthcare system’s resilience in the face of new public health challenges such as emerging infections is a key factor in achieving this goal [2]. This will necessitate continuous and sustained investments in capacity-building of both conventional and complementary healthcare systems such as community pharmacies to enhance their ability to adapt and respond effectively to the inevitable increases in demand for HBV services as countries work toward meeting the elimination targets for 2030.

Another notable finding of our study was that demand for HBV screening was lowest during the first quarter of each year, increasing linearly during subsequent quarters, and peaking in the last months of the year. This suggests the presence of seasonality and temporal patterns that could be further explored to optimize screening strategies and resource allocation for HBV care delivery in Sierra Leone. Studies from some settings have reported seasonality in viral hepatitis diagnosis while others have not. For example, in a study from the United Kingdom, Stewart et al [25] found a higher rate of HBV testing and diagnosis in the summer months, while Huang et al [26] reported higher rates of HBV diagnosis during spring and summer in China. Similarly, hepatitis C virus (HCV) diagnosis has been associated with the summer months in Mexico [27], Egypt [28], and China [26].

The reasons for seasonality in HBV and HCV diagnosis are not fully understood, however, since both are blood-borne pathogens, it has been suggested that seasonal variations in infections may be influenced by behavioral and climatic factors such as increased sexual activity, injection drug use, tattooing and travel to endemic areas during the summer months [25-29]. In areas with high HBV endemicity such as Sierra Leone, seasonal variations in HBV testing and diagnosis may be influenced by an entirely different set of factors such as cultural practices, differences in access to healthcare, healthcare policy and awareness campaigns. Low healthcare attendance in the first quarter may be attributable to the Christmas and New Year festivities in Sierra Leone. Of note, the KnowHep Foundation is heavily involved in awareness raising and outreach during the annual World Hepatitis Day in July, which could explain the increase in the utilization of our services observed in the third and fourth quarters. If this is the case, it would suggest that community-based advocacy and public awareness campaigns may successfully improve the frequency of screening in HBV-endemic areas in West Africa.

The overall prevalence of HBsAg was 17.5%, with the highest prevalence (24.4%) observed among individuals aged 30-39 years. This is in contrast to a recent systematic review and meta-analysis by Yendewa et al [19], which estimated a national HBsAg prevalence of 13.0% for Sierra Leone and a regional prevalence of 11.2% for the Western Area, where the present study was conducted. There are several possible explanations for this observed difference. Firstly, our pharmacy receives referrals from healthcare providers for clients with clinical suspicion of liver disease, many of whom are likely to have HBV infection. Secondly, our outreach and educational activities have attracted a substantial number of at-risk clients to our free or low-cost services, including those with family members or sexual partners with known HBV infection, those engaging in unprotected sexual activity, intravenous drug users, or those sharing personal hygiene items such as razors or toothbrushes. These groups have a higher pre-test probability of testing positive for HBV infection, which may partly explain the higher prevalence of HBsAg seropositivity observed in our study. Our findings are consistent with other studies that have also reported higher HBsAg seropositivity rates in healthcare settings, including hepatology clinics, sexually transmitted infection clinics, and hemodialysis centers [30].

These results must be interpreted within the limitations of study design. Firstly, due to a lack of record keeping, the pre-pandemic period was limited to one quarter, which may not accurately represent the baseline HBV testing trends. Secondly, the period over which trends were assessed is relatively short, further research is needed to confirm our conclusions over a more extended period. Thirdly, the study findings may not be generalizable to other HBV-endemic regions in Africa, given that the study was conducted in a single pharmacy in Freetown. Nonetheless, the study’s strengths lie in its contribution to the limited research on the impact of COVID-19 on HBV care delivery in SSA and its implications for the viral hepatitis elimination goals. Furthermore, our study showcases the critical, yet underappreciated role community pharmacies serve in resource-limited settings as primary access points for delivering HBV services to communities in need.

## CONCLUSION

In summary, the COVID-19 pandemic had a deleterious effect of HBV service delivery in a community pharmacy setting in Sierra Leone, with a 100% decrease in HBV screening observed during January to June of 2020 due to disruptions in global supply chains and population lockdowns. Notwithstanding, the total number of individuals screened increased modestly by 27% in the first year and by 23% in the second year after COVID-19-related population restrictions were eased, suggesting resilience in addressing short-term increases in demand for services. Over the three-year period of the study, HBV screening appeared to exhibit a seasonal trend, increasing linearly throughout the year. These findings may have implications for HBV elimination efforts in Sierra Leone and other highly endemic countries similarly affected by the COVID-19 pandemic.

## Data Availability

All data produced in the present study are available upon reasonable request to the authors

## AUTHOR CONTRIBUTIONS

GAY, MG, and RAS conceptualized and designed the study, with contributions from LSB, AMM, PBJ, SAY, SPEM, PEC, and SL. MG collected the data. GAY conducted the statistical analysis. All authors contributed to the interpretation of the data. MG, AMM and GAY wrote the initial manuscript draft. All authors critically reviewed the manuscript, contributed important intellectual content, and approved of the final version. GAY is acting as the guarantor of this manuscript.

## ACKNOWLEDGMENTS

We wish to acknowledge the pharmacy staff at CitiGlobe Ltd and people living with HBV in Sierra Leone, without whose help this study would not have been successful.

## DATA AVAILABILITY STATEMENT

The data presented in this study are available on request from the corresponding author upon reasonable request.

## ETHICAL APPROVAL

Ethical approval to perform the study was obtained from Njala University, Bo Campus, Sierra Leone (approval date 23 January 2023). Informed consent was not required as this study involved a retrospective analysis of secondary data.

